# Cross-country generalizability of foundation models for cervical cancer screenings on H&E whole slide images

**DOI:** 10.64898/2026.07.22.26358575

**Authors:** Martin Paulikat, Christoph Bosch, Witali Aswolinskiy, Isabella Caixeta Borges, Lennart Nauschütte, Christian Aichmüller, Dietmar Schmidt, Hermann Bussmann, Simon Kalteis, Miroslav Zapukhlyak, Magnus von Knebel Doeberitz, Matthias Kloor

## Abstract

Accurate grading of cervical biopsies on Hematoxylin and Eosin (H&E) stained whole slide images (WSIs) is essential for distinguishing high grade lesions from low grade changes, yet this process is subject to considerable inter-observer variability. In this study, we evaluate a foundation model-based multiple instance learning (MIL) pipeline for binary high-grade squamous intraepithelial lesion (HSIL) detection on H&E stained WSIs. We benchmark our Athena foundation model against four state-of-the-art pathology foundation models: H-optimus-0, Hibou-L, Midnight-12k and Virchow, across datasets from five different countries: Portugal, Cambodia, Germany, Poland and Scotland. Athena achieved the highest mean area under the curve (AUC) (0.931) with the lowest cross-country variability (STD = 0.022). Furthermore, we compared the model’s diagnostic performance to that of trained pathologists on a dataset with p16-confirmed ground truth. Our model improved sensitivity from 84% to 95% while maintaining comparable specificity (85% vs. 84%). Failure analysis revealed that the model’s errors were concentrated at the diagnostic boundary between low-grade and high-grade lesions, whereas pathologists’ errors spanned a broader range of misclassifications. These findings show the potential of foundation models for cervical cancer screenings worldwide.

## 1 Introduction

Cervical cancer remains one of the leading causes of cancer-related mortality among women worldwide, particularly in low- and middle-income countries (LMICs). Persistent infection with high-risk human papillomavirus (HPV) can progress through multiple precancerous stages. Most critical is the stage of high-grade squamous intraepithelial lesion (HSIL), which is highly treatable when detected early, but has a high risk of turning into an invasive carcinoma.

The diagnosis of cervical precancerous lesions follows a multi-step process. Primary screening tools include high-risk HPV DNA testing, cytology approaches, like the Papanicolaou (Pap) test and visual inspection with acetic acid without magnification (VIA) which is mostly used in low resource settings. Abnormal cytology results or detection of high risk HPV are further evaluated by colposcopy, during which the cervix is examined under magnification to identify suspicious areas. When colposcopic findings suggest the presence of a lesion, biopsies are obtained for histopathological examination.

Histopathological assessment represents the gold standard for grading cervical precancerous lesions. They are traditionally classified by the cervical intraepithelial neoplasia (CIN) grading system, which distinguishes between Normal, CIN1, CIN2, CIN3 and invasive cancer based on the progression of transforming HPV infection in the epithelium. More recently, the WHO has adopted a two-tiered classification that groups lesions into low-grade squamous intraepithelial lesion (LSIL), corresponding to CIN1 and HSIL, encompassing CIN2 and CIN3 (Herrington et al., 2020). This distinction is of direct clinical relevance, as LSIL typically reflects a transient HPV infection with high rates of spontaneous regression, whereas HSIL carries a significant risk of progression to invasive cervical carcinoma if left untreated. Therefore the histopathological assessment serves as the critical decision point for further treatment recommendation.

Accurate histological grading is essential, as misclassification has direct consequences for patient outcomes. Over-diagnosis of LSIL as HSIL can lead to unnecessary invasive procedures such as loop electrosurgical excision procedure (LEEP) or conization, which carry risks including cervical stenosis, hemorrhage, and adverse obstetric outcomes in pregnancy for women of reproductive age (Pina, Lavallée, Ndiaye, & Mayrand, 2013). Conversely, under-diagnosis of HSIL may result in delayed treatment, allowing precancerous changes to progress to invasive carcinoma.

Traditionally, cervical biopsy specimens are evaluated using Hematoxylin and Eosin (H&E) staining, with histopathological interpretation performed by expert pathologists. Immunohistochemical (IHC) staining for p16 is widely used as an adjunctive biomarker to improve the accuracy and reproducibility of H&E-based diagnoses, particularly in diagnostically challenging cases. However, p16 IHC requires specialized laboratory infrastructure, and technical expertise that are frequently unavailable outside well-resourced pathology departments, meaning that in many LMICs, biopsies must be assessed on H&E morphology alone, without the diagnostic support that p16 confirmation provides in high-income settings. This limitation is further compounded by a shortage of trained pathologists in many of these regions: many sub-saharan countries only have on average 1 pathologist per 1,000,000 inhabitants, while in the US and UK the rate of pathologists per inhabitant lies between 1 in 20,000 (Mudenda, Malyangu, Sayed, & Fleming, 2020; Nelson, Milner, Rebbeck, & Iliyasu, 2016). As a result, biopsies in these settings may be evaluated by general pathologists or even non-specialist clinicians, based on H&E morphology alone and without access to IHC confirmation. A diagnostic tool that performs reliably on H&E-stained slides alone, regardless of the clinical setting in which it is deployed, could therefore help compensate for the unavailability of p16 IHC and would be of considerable value in reducing these global inequities in cervical cancer care.

The digitization of histological slides into whole slide images (WSIs) has opened new opportunities. WSIs capture the entire tissue section at high resolution, typically at 20× magnification, resulting in large gigapixel images. The sheer size of WSI poses significant computational challenges, as they exceed the memory capacity of current GPU hardware when loaded in their entirety and therefore prohibit direct processing by classification models. A common strategy is therefore to partition each WSI into thousands of smaller tiles or patches, which can be individually processed. However, this introduces a second challenge: diagnostic labels are typically available only at the slide level while the precise location of the lesion within the slide is often not annotated. This weakly supervised setting, in which a single label must be inferred from a large collection of unlabeled instances, is naturally formulated as a multiple instance learning (MIL) problem.

Recently, foundation models have gained attention in digital pathology. These methods use large pretrained models to compress patches of WSIs into high-level feature representations that are aggregated for slide-level classification. They have demonstrated strong performance in other histopathology tasks (Neidlinger et al., 2025). In the context of cervical cancer, (Aswolinskiy, van der Post, et al., 2025) evaluated multiple pretrained encoders within an attention MIL framework for multi-class cervical lesion classification. (Li, Feng, Wang, & Xu, 2021) applied a similar MIL strategy in the context of the TissueNet challenge (DrivenData, 2020), achieving competitive but not leading performance in the challenge. (Zhao et al., 2022) employed a masked autoencoder to learn WSI representations in a self-supervised manner before classification. Alternative strategies include segmentation approaches to elaborate the HPV progression (An et al., 2023; Andreassen, Mortensen, Stenbro, Sørensen, & Sørbye, 2025; Mohammadi et al., 2024; Oliveira et al., 2023; Sornapudi et al., 2021), modeling the spatial relationship between neighboring patches (Wang et al., 2023) and confidence-based patch selection (Jiang et al., 2022), while others rely on handpicked patch selection before analysis (Cho et al., 2022; Habtemariam, Zewde, & Simegn, 2022; Pal et al., 2021).

We address these gaps by benchmarking our Athena foundation model (Bosch et al., 2026) in the MIL setting against current state-of-the-art pathology foundation models for HSIL detection in H&E-stained WSIs. While H-optimus-0 and Hibou-L do not disclose country-diversity, Virchow only used data from a single center and Midnight-12k used the TCGA database which is primarily dominated by US data (Karasikov et al., 2025; Nechaev, Pchelnikov, & Ivanova, 2024; Saillard et al., 2024; Vorontsov et al., 2024; Weinstein et al., 2013). In contrast, Athena was explicitly built to prioritize diversity over sheer quantity. Here, diversity encompasses not only geographic and demographic variation across patient populations, but also technical variation such as differing staining protocols and scanner hardware. Despite being trained on only 274,150 WSIs, fewer than Virchow (∼ 1,5M), Hibou-L (∼ 1M), or H-optimus-0 (∼ 500k), its training data spanned 52 centers across 25 countries and 8 different scanners. Evaluating on datasets from five different countries, we show that our approach generalizes across diverse clinical settings and provides a viable path toward equitable cervical cancer diagnostics worldwide. Additionally, we compared the results of our model directly to those of trained pathologists on the Cambodian dataset, showing that it could improve diagnostic performance in LMIC settings.

## 2 Materials and Methods

This section describes the datasets used in this study, followed by the preprocessing pipeline, an introduction to foundation and MIL models, experimental setup and finally the evaluation criteria.

### 2.1 Datasets

For this study, we used three publicly available and two proprietary datasets from five countries: Portugal, Cambodia, Germany, Poland, and Scotland. All datasets are summarized in Table 1.

**Table 1:**
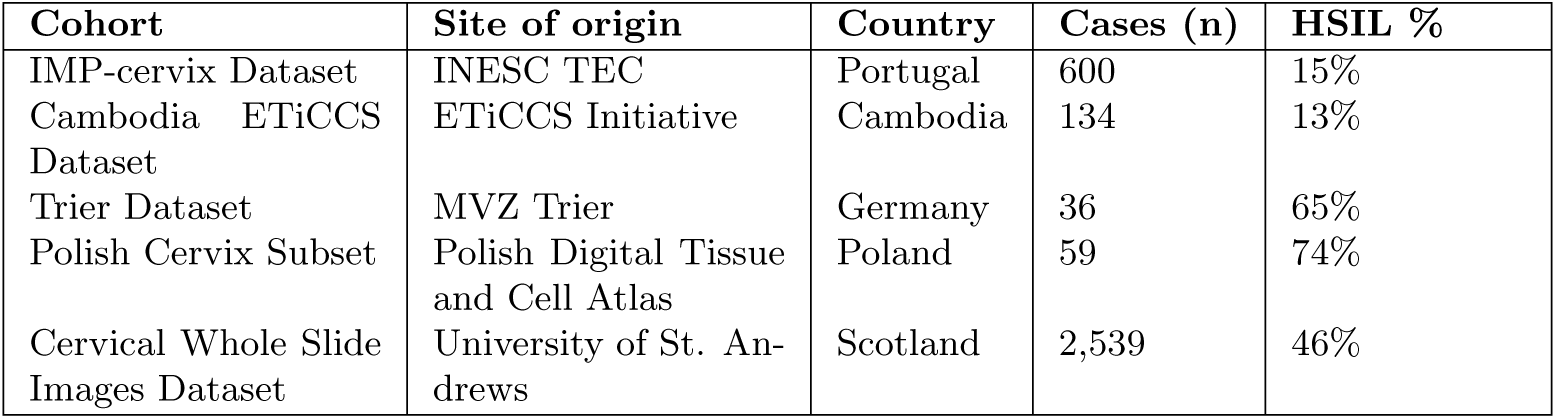
Overview of the five cervical cancer histopathology datasets used in this study. Datasets originate from five countries across Europe and Southeast Asia, representing diverse clinical and technical settings, patient populations. HSIL prevalence ranges from 13% to 74%.

#### IMP Cervix Dataset

Extracted from the data archive of the IMP diagnostic laboratory, part of INESC TEC in Porto, Portugal. The dataset contains 600 cervical LEEP samples, with one image per patient. The HSIL rate is 15%. This dataset served as the training set for all classification models. The dataset is publicly available (Oliveira et al., 2023).

#### Cambodia ETiCCS Dataset

Collected as part of a cervical cancer study in Cambodia by the ETiCCS Initiative. The dataset comprises 134 cases with one image per patient and an HSIL rate of 13%. All biopsies were initially evaluated on H&E staining by trained pathologists in Cambodia and subsequently reviewed with p16 IHC staining by a senior pathologist in Germany, whose assessment served as the ground truth. This dataset was used for testing and for comparing the performance of our Athena-based MIL model with that of the Cambodian pathologists. The dataset is proprietary.

#### Trier Dataset

Extracted from the archive of MVZ Trier in Germany. The dataset contains 36 cases, with some cases having more than one image per patient, leading to 185 images. The HSIL rate is 65%. The dataset was used for testing. The dataset is proprietary.

#### Polish Cervix Subset

Extracted from the Polish Digital Tissue and Cell Atlas, containing 59 cases. Some cases have more than one image per patient, leading to 401 images. The HSIL rate is 74%. The dataset was used for testing and is publicly available (Skokowski et al., 2022).

#### Cervical Whole Slide Images Dataset

2,539 images from the University of St. Andrews in Scotland, UK. Each case contains one image per patient. The HSIL rate is 46%. The dataset was used for testing and is publicly available (Mohammadi et al., 2025).

### 2.2 Preprocessing

Preprocessing consisted of three steps. First, tissue area was identified in each WSI using thresholding in the color space. Each slide was transformed into the CIELAB color space, and regions exceeding a threshold of 135 in the *a^∗^* channel were classified as tissue. To remove small and noisy content, small holes and isolated objects were removed using morphological filtering (van der Walt et al., 2014), resulting in a binary tissue mask. An example of the LAB-based tissue detection is shown in Figure 1. WSIs are typically stored as multi-resolution pyramidal TIFF files, which have multiple levels of spatial resolution. The resolution is commonly expressed as microns (*µ*m) per pixel, where lower values correspond to higher magnification. From the detected tissue regions, non-overlapping patches of 224 × 224 pixels were extracted at spacing 1 (*µ*m) per pixel. Finally, each patch was normalized using the mean and standard deviation values corresponding to the respective foundation model pretraining configuration. These patches served as input to the pretrained foundation models described in the following subsection.

**Figure 1:**
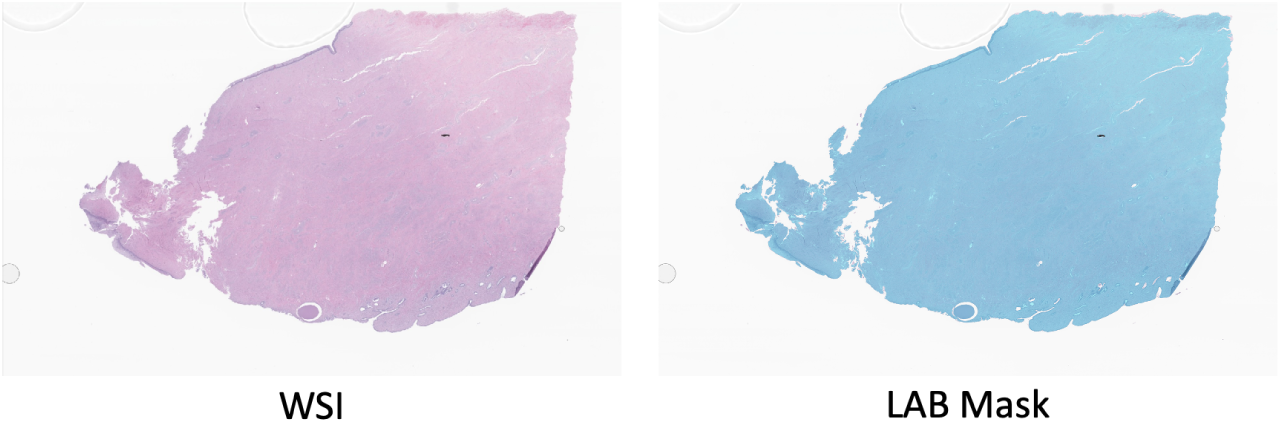
Example of CIELAB color space tissue detection. Left: original H&E-stained WSI. Right: corresponding binary tissue mask overlaid on the original image. Blue areas indicate detected tissue content.

### 2.3 Foundation Models

WSIs pose a significant computational challenge due to their large size. Processing an entire WSI at once with a classification model is infeasible, as it far exceeds the memory capacity. While processing slides at lower resolutions would reduce the memory requirements, clinically important details could be lost. For cancer subtyping tasks, spacings at 0.5*µ*m per pixel or 1.0*µ*m per pixel are commonly used. At a spacing of 1.0*µ*m per pixel, a single WSI from the Portuguese dataset has a mean resolution of 10.070 × 32.666 pixels with 3 color channels (rgb), making direct processing impossible.

Foundation models address this problem by compressing tissue patches into compact feature representations. In this paradigm, a WSI is partitioned into thousands of smaller patches *p* after background removal, each of which is compressed by the pretrained foundation model into feature vectors of the size *d*. The compress factor *cf* can therefore be shown as:

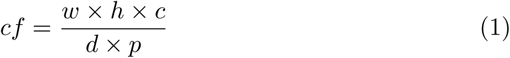

where *w* and *h* are the pixel width and height of the uncompressed image at a chosen *µ*m per pixel and *c* is the number of color channels. Using the values of the aforementioned example, the Athena model achieves a compress factor of:

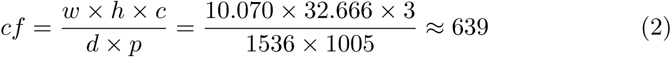

as on average 1,005 patches were extracted per image and each patch was transformed into a feature vector of size 1,536.

Foundation models are typically based on vision transformers (Zhai, Kolesnikov, Houlsby, & Beyer, 2022), and pretrained on large datasets of histological images using self-supervised learning. These models learn general-purpose histological representations that can effectively be used in a wide range of downstream tasks without requiring task-specific retraining.

In this work we evaluated our own foundation model, Athena (Bosch et al., 2026), alongside four publicly available alternatives: H-optimus-0 (Saillard et al., 2024), Hibou-L (Nechaev et al., 2024), Midnight-12k (Karasikov et al., 2025), and Virchow (Vorontsov et al., 2024).

### 2.4 Multiple Instance Learning

After compression by the foundation model, the resulting feature vectors are aggregated and processed by a classifier. A common method for this task is attention-based MIL, as it is well suited for slide level prediction in absence of patch level labels (Ilse, Tomczak, & Welling, 2018). The MIL classifier consists of two components: an attention mechanism that learns to identify the most diagnostically relevant compressed patches within each WSI, and a classification head that aggregates the attended features to produce a slide level prediction. In our study, the output is a binary classification score indicating the likelihood that the processed WSI contains HSIL regions. The complete pipeline is shown in Figure 2.

**Figure 2:**
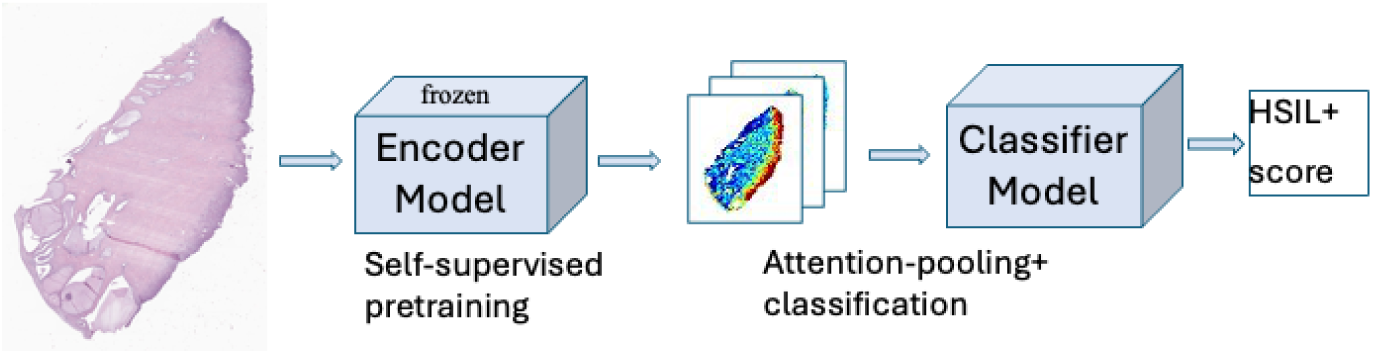
Pipeline from feature extraction by the foundation model to down-stream processing in the MIL classification model.

### 2.5 Experimental Setup

The Portuguese dataset was used as the training set for all experiments. We deliberately trained on a single dataset in order to measure out-of-distribution performance on the external test datasets. Single-cohort training suffices because the MIL classifier operates on frozen foundation model features, so generalization derives from the foundation model’s pretraining. Tissue regions were detected using the LAB color space method and subsequently compressed into feature representations by each respective foundation model. We conducted experiments using five foundation models: Athena, H-optimus-0, Hibou-L, Midnight-12k, and Virchow. For each model, a binary HSIL MIL classification ensemble was trained using five-fold cross-validation on the Portuguese dataset. The resulting ensemble was evaluated on a held-out Portuguese test set as well as on the Cambodian, German, Polish, and Scottish datasets.

The MIL classifier model was trained using the Adam Optimizer (Kingma & Ba, 2014) with an initial learning rate of 0.001 and cross-entropy loss. A ReduceLROnPlateau scheduler (factor 0.1, patience 3) was applied to adjust the learning rate based on validation loss. Training was performed for a maximum of 100 epochs per fold with early stopping (patience = 30) based on validation area under the curve (AUC). Each experiment was repeated across six random seeds, and we report the mean AUC performance across runs. All experiments were conducted on a single NVIDIA L4 GPU using PyTorch.

### 2.6 Evaluation

We evaluated our approach using three analyses:

#### Performance evaluation

We computed the AUC, sensitivity and specificity of the Athena-based MIL classifier on the held-out Portuguese and each other test set. As the Polish and German datasets contain multiple images per patient, we used the highest predicted score per patient as the patient-level prediction.

#### Cross-country comparison

We compared the Athena foundation model against four alternative foundation models on all test datasets. For each model we report the AUC per dataset as well as the mean AUC and standard deviation across all datasets. Differences between Receiver Operating Characteristic (ROC) curves were assessed using DeLong’s test, with *p* ≤ 0.05 considered statistically significant.

#### Comparison with Cambodian Pathologists

We evaluated our model’s performance with that of the Cambodian Pathologists, who graded the cases in the Cambodian dataset, comparing sensitivity and specificity. The ground truth was determined by p16 IHC staining performed by a senior pathologist in Germany. We additionally conducted a failure analysis examining all false positives and false negatives produced by both the pathologists and our model.

## 3 Results

In this section, we first present a comprehensive performance analysis of our model. We then benchmark our approach against state-of-the-art pathology foundation models across datasets from five different countries. Finally, we compare the model’s performance to that of local pathologists on the Cambodian dataset.

### 3.1 Athena Performance

We evaluated the Athena-based MIL classifier on the held-out Portuguese test set and on all other test datasets. For each dataset, we computed the AUC, sensitivity and specificity. As shown in Table 3, our Athena-based MIL model achieved strong performance across all five datasets, with AUCs ranging from 0.910 on the German dataset to 0.963 on the Cambodian dataset. Sensitivity ranged from 78% on the Portuguese dataset to 95% on the Cambodian dataset, while specificity ranged from 77% on the German dataset to 93% on the Portuguese dataset. The corresponding confusion matrices for each dataset are shown in Table 2. Additionally, we show two examples of attention maps in Figure 3. The model’s attention focuses on the epithelium area which is the expected site of lesion development.

**Figure 3:**
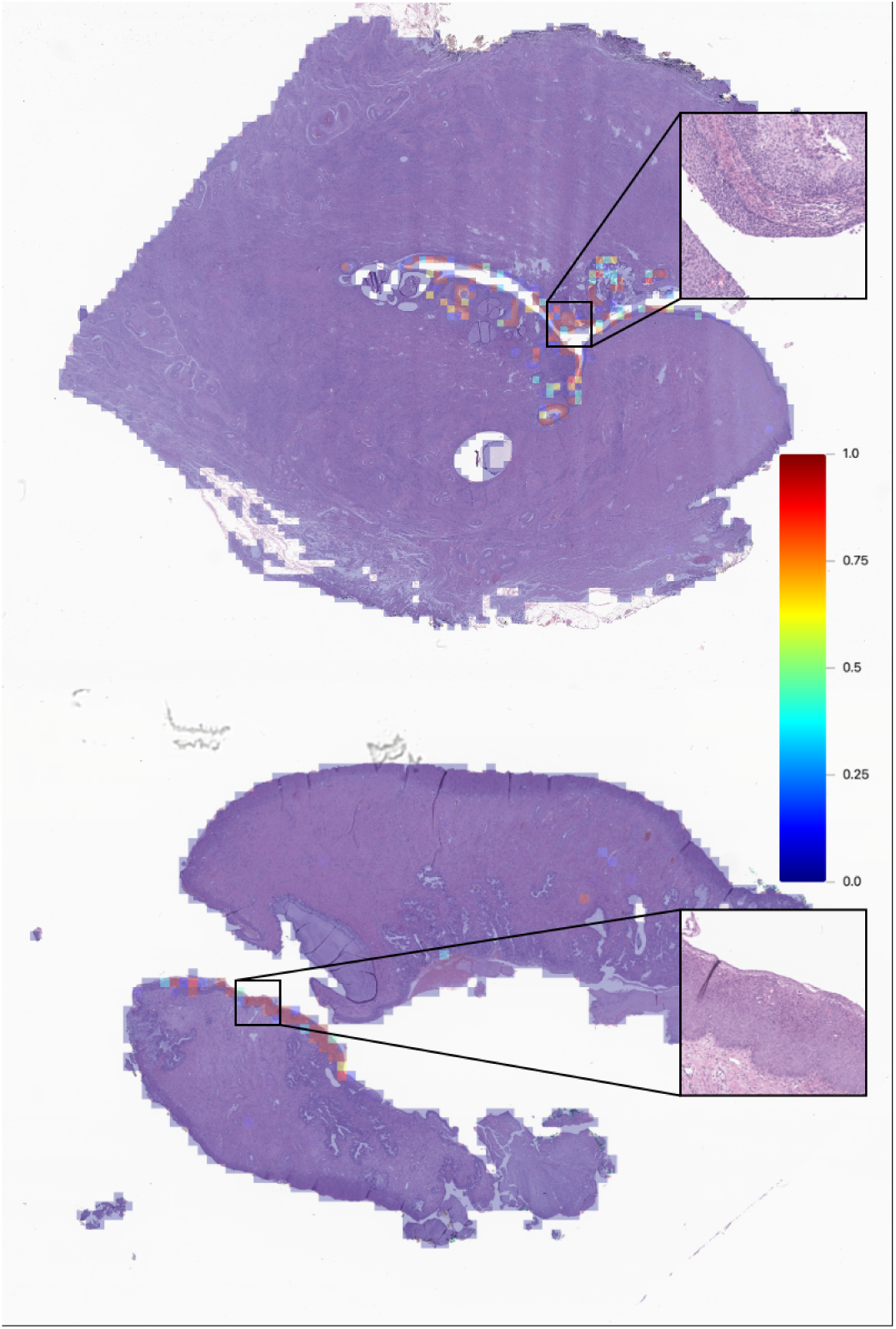
Two attention map examples from the Portuguese test dataset. Both images were correctly classified as HSIL using feature vectors from the Athena foundation model. The model’s attention concentrates on the epithelial region, which is consistent with the expected site of CIN lesions, as shown in as shown in the zoomed-in H&E tiles, which highlights the dysplastic squamous epithelium.

**Table 2:**
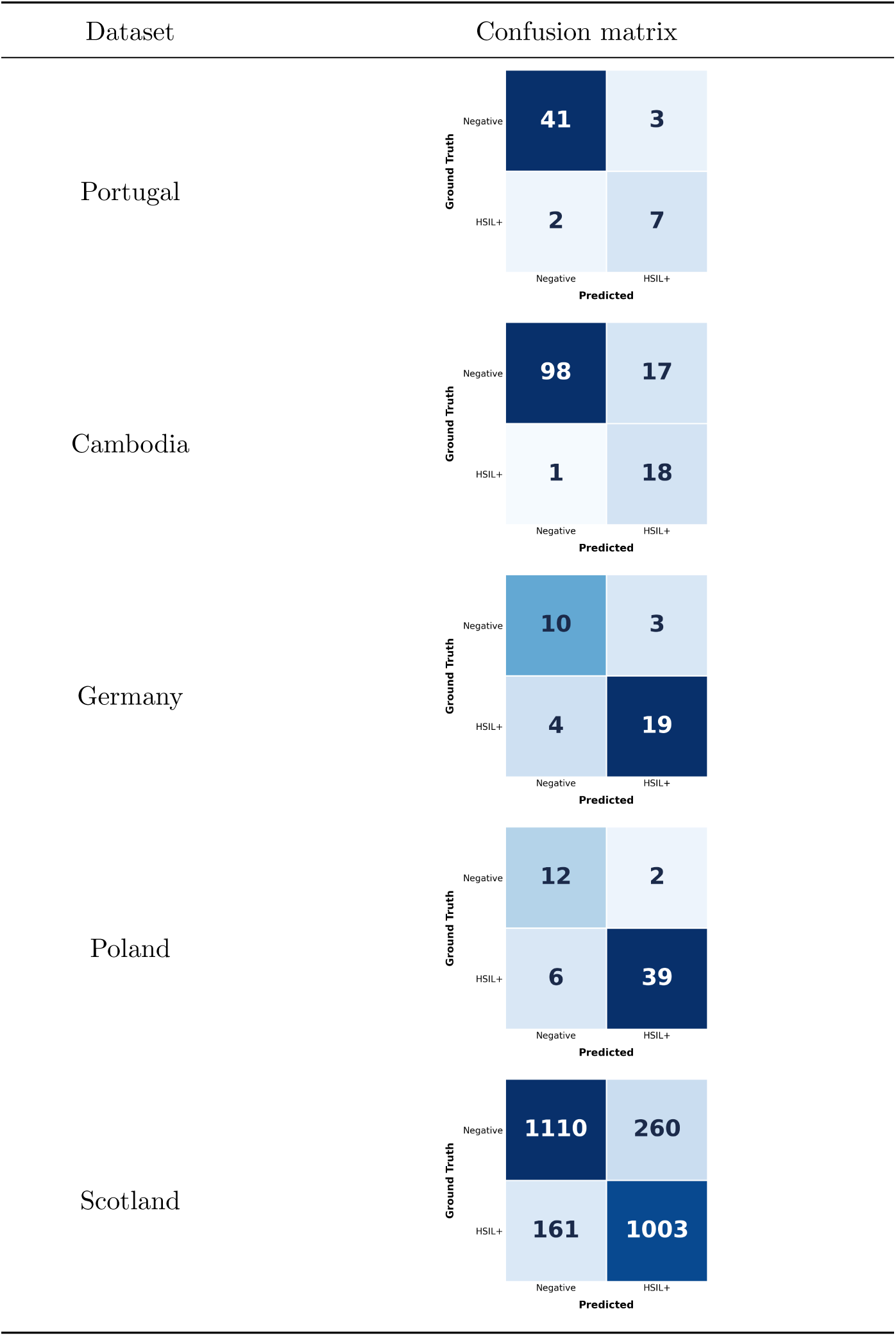
Confusion matrices for each dataset, showing the predictions of the Athena foundation model-based MIL classifier.

**Table 3:** Performance of the Athena-based MIL classifier on each dataset in terms of sensitivity, specificity, and AUC.

| Dataset | Sensitivity | Specificity | AUC |
| --- | --- | --- | --- |
| Portugal | 78% | 93% | 0.942 |
| Cambodia | 95% | 85% | 0.963 |
| Germany | 83% | 77% | 0.910 |
| Poland | 87% | 86% | 0.925 |
| Scotland | 83% | 86% | 0.913 |

### 3.2 Comparison of Foundation Models across Countries

We benchmarked the Athena foundation model against four alternative foundation models: H-optimus-0, Hibou-L, Midnight-12k, and Virchow on all test datasets. As described in Section 2.6, all models were trained on the Portuguese dataset and evaluated on a held-out Portuguese test set as well as on all external datasets.

The results are shown in Table 4 and visualized in Figure 4. Athena achieved the highest mean AUC across all datasets (0.931) with the lowest standard deviation (0.022). Athena achieved the highest performance for the Cambodian (0.963), Polish (0.925) and Scottish dataset (0.913), while H-optimus-0 obtained the highest AUC on the Portuguese (0.955) and the German dataset (0.923).

**Figure 4:**
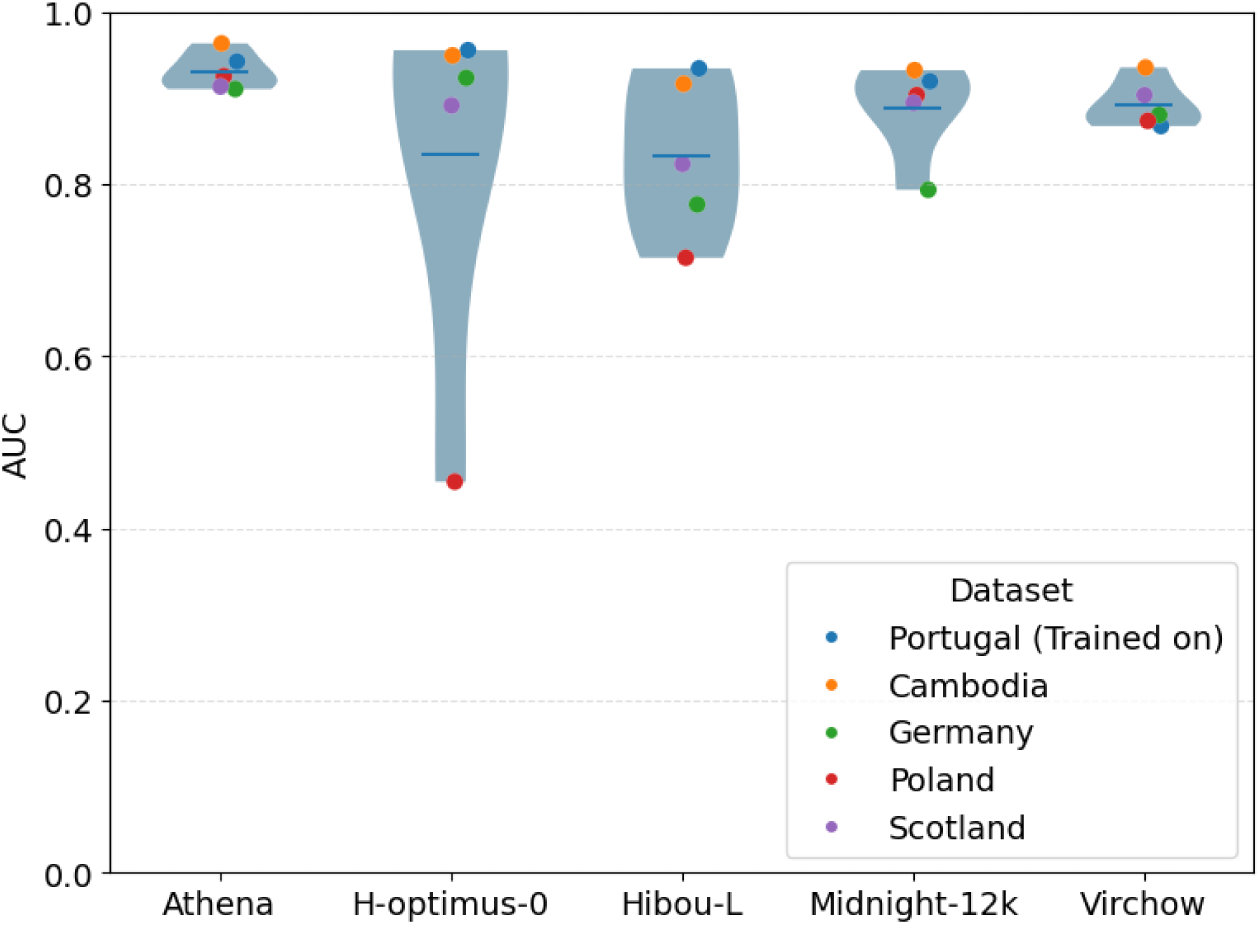
Performance comparison of the Athena foundation model against state-of-the-art pathology foundation models on the downstream HSIL detection task. All classification models were trained on the Portuguese dataset and evaluated on a held-out test set from the same cohort, as well as datasets from Cambodia, Germany, Poland and Scotland.

**Table 4:** AUC scores for HSIL detection across five country-specific datasets, comparing the Athena foundation model against four state-of-the-art pathology foundation models. All models were trained on the Portuguese dataset and evaluated on a held-out Portuguese test set as well as datasets from Cambodia, Germany, Poland, and Scotland. Bold values indicate the highest AUC per dataset. Athena achieves the highest mean AUC (0.931) with the lowest variability across sites (STD = 0.022).

|  | Athena | H-optimus-0 | Hibou-L | Midnight-12k | Virchow |
| --- | --- | --- | --- | --- | --- |
| Portugal | 0.942 | **0.955** | 0.934 | 0.919 | 0.867 |
| Cambodia | **0.963** | 0.949 | 0.916 | 0.932 | 0.935 |
| Germany | 0.910 | **0.923** | 0.776 | 0.793 | 0.880 |
| Poland | **0.925** | 0.454 | 0.714 | 0.903 | 0.873 |
| Scotland | **0.913** | 0.891 | 0.823 | 0.894 | 0.903 |
| Mean | **0.931** | 0.834 | 0.833 | 0.888 | 0.892 |
| STD | **0.022** | 0.214 | 0.093 | 0.055 | 0.028 |

To assess whether these differences were statistically significant, we compared the ROC curves of Athena against each competing model using DeLong’s test (Supplementary Table S1). Athena’s AUC was significantly higher than that of H-optimus-0 and Hibou-L on the Polish and Scottish datasets, and than Midnight-12k on the Cambodian and Scottish datasets, while differences on the remaining comparisons did not reach significance.

### 3.3 Comparison with Pathologist Performance on the Cambodian Dataset

To contextualize our model’s diagnostic accuracy relative to human experience in a real-world clinical setting, we compared its predictions to those of experienced Cambodian pathologists who evaluated the Cambodian ETiCCS dataset. All cases were confirmed using p16 IHC staining by a senior pathologist in Germany, whose assessment served as ground truth. The results are summarized in Table 5.

**Table 5:**
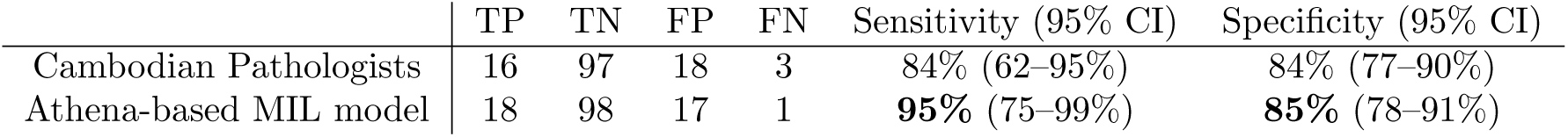
Comparison of HSIL detection performance on the Cambodian dataset (n = 134) between local Cambodian pathologists and our foundation model MIL approach. Ground truth was obtained by a senior German pathologist with p16 IHC staining, who detected 19 HSIL and 115 negative cases. The Cambodian pathologists achieved a sensitivity of 84%, with a CI of 62-95%. They over-diagnosed 18 cases as HSIL, resulting in a specificity of 84% and a CI of 77-90%. Our model missed only one positive case, increasing the sensitivity to 95%, with a CI between 75-99% and reduced the number of false positives by 1, leading to a specificity of 85%, with a CI of 78-91%.

|  | TP | TN | FP | FN | Sensitivity (95% CI) | Specificity (95% CI) |
| --- | --- | --- | --- | --- | --- | --- |
| Cambodian Pathologists | 16 | 97 | 18 | 3 | 84% (62–95%) | 84% (77–90%) |
| Athena-based MIL model | 18 | 98 | 17 | 1 | **95%** (75–99%) | **85%** (78–91%) |

Of the 19 HSIL cases, 16 were correctly detected by the Cambodian pathologists, resulting in a sensitivity of 84%, with a confidence interval (CI) of 62-95%. The three missed cases comprised one adenocarcinoma and two CIN2 lesions. Additionally, 18 of the 115 negative cases were falsely classified as HSIL, resulting in a specificity of 84%, with a CI of 77-90%. Among the over-diagnosed cases, 11 were graded as CIN2, two as CIN3, three as being between CIN2 and CIN3 and two as carcinomas, of which 17 were confirmed as being metaplasia and one as CIN1.

In comparison, our model achieved a sensitivity of 95%, with a CI between 75-99% and a specificity of 85%, with a CI of 78-91%, on the same dataset (AUC = 0.963), with 17 false positives and only one false negative. The single missed HSIL case was a CIN2 case. Among the false positives, the model over-diagnosed 10 metaplasia, one cervical polyp and six CIN1 cases. From the six over-diagnosed CIN1 cases two have been confirmed as being p16 positive.

## 4 Discussion

In this study we investigated whether foundation and MIL models can provide competitive and generalizable HSIL detection in H&E stained cervical cancer WSIs. We addressed two key questions: first, whether foundation models generalize well over different diverse populations and clinical settings, and second, how such models compare to the diagnostic performance of pathologists in LMIC settings.

Our results demonstrate that the Athena foundation model, combined with an attention-based MIL classifier, achieves strong HSIL detection performance across all five datasets, with AUCs ranging from 0.910 to 0.963. The cross-country benchmarking revealed that Athena achieved the highest mean AUC (0.931) with the lowest variability across sites (STD = 0.022), outperforming H-optimus-0, Hibou-L, Midnight-12k, and Virchow. Notably, H-optimus-0 surpassed Athena on the Portuguese dataset which was used for training the MIL classifier models, but in general performed worse across all countries, underscoring that single-site performance is an insufficient proxy for real-world applicability and that cross-country validation is essential. The superior generalizability of Athena may be attributed to its pretraining strategy, which prioritized data diversity over sheer quantity, as it was trained on data from 25 different countries (Bosch et al., 2026). This has direct implications for model selection in global health settings, where training and deployment conditions are rarely matched.

Notably, the five datasets varied considerably in HSIL prevalence, ranging from 13% in the Cambodian cohort to 74% in the Polish cohort. Such differences likely reflect not only variation in underlying HPV prevalence across populations, but also differences in the sensitivity and coverage of upstream screening programs that determine which patients are referred to colposcopy and biopsy in the first place. Despite this substantial variation in case-mix, Athena maintained consistently high and stable performance across all cohorts. This robustness to prevalence shifts is an important property for real-world deployment, since screening infrastructure and referral pathways differ widely between countries, and a model’s performance in a single setting is not necessarily representative of the population it would encounter elsewhere.

Compared to local pathologists on the Cambodian dataset, our model improved sensitivity from 84% to 95% while maintaining comparable specificity (85% vs. 84%). Error patterns differed: pathologists predominantly over-graded metaplasia cases (Tranbaloc, 2002), while the model’s errors were concentrated at the LSIL/HSIL boundary, suggesting that computational and human assessment may be complementary. A limitation of the pathologist comparison is the small size of the p16-confirmed HSIL group (n = 19), which yields wide confidence intervals around the sensitivity estimates. Larger cohorts are needed to establish whether the observed sensitivity advantage is robust.

Future work of this study will include prospective clinical studies to evaluate whether the model can be integrated into screening workflows to support pathologists in LMICs. Expanding the evaluation to additional countries, particularly in Sub-Saharan Africa and South Asia, would consolidate our generalizability claims. Additionally, incorporating explainability methods such as lesion segmentation maps facilitate clinical adoption by allowing pathologists to verify the model’s diagnostic reasoning.

To make our approach viable in LMICs, it could be deployed via a cloud-based system, similar to the one described by (Aswolinskiy, Motiwala, et al., 2025) for MSI/MSS classification in colorectal cancer. Slides would be scanned locally using a whole slide scanner, uploaded to a remote server running the model, and returned as classification results. Using this system there will be no need for high-computing hardware on-site. Furthermore, the system could be integrated within electronic health information systems used e.g. in cancer screening. Classification results would then be automatically linked to patients’ electronic health records, making it easier to track lesion progression and facilitate the comparison with results of complementary tests such as HPV testing.

Cervical cancer remains a global health challenge that disproportionately affects regions with the least access to diagnostic expertise. In this study, we demonstrated that foundation model-based MIL can confidently detect HSIL lesions with high accuracy and that this performance generalizes across geographically diverse datasets when it was explicitly prioritized in the foundation model pretraining. In addition, we showed that foundation model-based MIL approaches reduce diagnostic variability and improve the performance in direct comparison with pathologists. Foundation models may play an important role for an equitable cervical cancer diagnostic worldwide.

## 5 Supplementary

**Table S1:** Statistical tests using the DeLong’s test. For the tests we compare the prediction results of our Athena-based MIL method against four other state-of-the-art pathology foundation models and reported the p-values. Significance was found for p-values ≤ 0.05 and colored in green, while insignificance was colored in red.

|  | Athena vs.<br>H-optimus-0 | Athena vs.<br>Hibou-L | Athena vs.<br>Midnight-12k | Athena vs.<br>Virchow |
| --- | --- | --- | --- | --- |
| Portugal | 0.582 | 0.781 | 0.578 | 0.160 |
| Cambodia | 0.189 | 0.157 | 0.039 | 0.273 |
| Germany | 0.779 | 0.090 | 0.057 | 0.590 |
| Poland | <0.001 | 0.010 | 0.568 | 0.267 |
| Scotland | <0.001 | <0.001 | <0.001 | 0.075 |

## Competing Interests

C.A. is a founder of PAICON GmbH. C.B., W.A., S.K., and M.Z. are employees of PAICON GmbH. M.v.K.D. is affiliated with MoCaP GmbH. D.S. is affiliated with MVZ Trier. All other authors declare no competing interests.

## Data Availability

Three of the five datasets analyzed in this study are publicly available. The IMP-cervix dataset is available from Oliveira et al. (2023): https://rdm.inesctec.pt/dataset/nis-2023-008. The Polish Cervix Subset is available as part of the Polish Digital Tissue and Cell Atlas (Skokowski et al., 2022): https://mostwiedzy.pl/en/publication/the-digital-tissue-and-cell-atlas-and-the-virtual-microscope,158247-1. The Scottish Cervical Whole Slide Images dataset is available from Mohammadi et al. (2025): https://zenodo.org/ records/10276838. The Cambodia ETiCCS dataset and the Trier dataset are not publicly available. These datasets may be made available from the corresponding author on reasonable request.

## Author Contributions

M.P. designed and implemented the study, conducted the experiments and analyses, and wrote the manuscript. C.B. developed the Athena foundation model. W.A., C.A. and M.v.K.D. provided scientific advice. I.C.B. and L.N. contributed medical expertise. S.K. and M.Z. contributed to data acquisition and preparation. D.S. and H.B. provided datasets. M.K. supervised the study. All authors read and approved the final manuscript.

## Notes

### Author Declarations

This study is a retrospective analysis of pseudonymized whole-slide images from multiple cervical cancer cohorts. All data was collected in accordance with local ethical and legal requirements and anonymized prior to analysis. Ethical approval was obtained from the National Ethics Committee for Health Research (NECHR) (NECHR 058), Cambodia and the Ethics Commission of the Heidelberg University School of Medicine, Germany (S-136/2022). All participants provided signature or thumbprint on informed consent form in Khmer before enrollment; other sites obtained equivalent local approvals. External data was processed on-site at the corresponding institutions.

## References

An, H., Ding, L., Ma, M., Huang, A., Gan, Y., Sheng, D., . . . Zhang, X. (2023). Deep learning-based recognition of cervical squamous interepithelial lesions. Diagnostics, 13 (10), 1720.

Andreassen, A. K., Mortensen, E., Stenbro, R., Sørensen, Ø., & Sørbye, S. W. (2025). Digital pathology with ai for cervical biopsies: Diagnostic accuracy at the cin2+ threshold. Cancers, 17 (23), 3808.

Aswolinskiy, W., Motiwala, M., Paulikat, M., Nelius, N., Ahadova, A., Adam, P., . . . Aichmüller, C. (2025). Uncertainty-aware prediction of microsatellite instability in colorectal cancer from h&e-stained whole slide images. medRxiv, 2025–11.

Aswolinskiy, W., van der Post, R. S., Campora, M., Baronchelli, C., Ardighieri, L., Vatrano, S., . . . others (2025). Attention-based whole-slide image compression achieves pathologist-level pre-screening of multi-organ routine histopathology biopsies. Modern Pathology, 100827.

Bosch, C., Wong, J. K., Paulikat, M., Zapukhlyak, M., Arora, B., Aichmüller-Ratnaparkhe, M., . . . others (2026). Diversity over scale: Whole-slide image variety enables h&e foundation model training with fewer patches. Journal of Pathology Informatics, 100648.

Cho, B.-J., Kim, J.-W., Park, J., Kwon, G.-Y., Hong, M., Jang, S.-H., . . . Park, S.-T. (2022). Automated diagnosis of cervical intraepithelial neoplasia in histology images via deep learning. Diagnostics, 12 (2), 548.

DrivenData. (2020). Tissuenet: Detect lesions in cervical biopsies. https://www.drivendata.org/competitions/67/competition-cervicalbiopsy/page/254/. (Online; accessed 10 December 2025)

Habtemariam, L. W., Zewde, E. T., & Simegn, G. L. (2022). Cervix type and cervical cancer classification system using deep learning techniques. Medical devices: evidence and research, 163–176.

Herrington, C. S., et al. (2020). Who classification of tumours female genital tumours.

Ilse, M., Tomczak, J., & Welling, M. (2018). Attention-based deep multiple instance learning. In International conference on machine learning (pp. 2127–2136).

Jiang, P., Liu, J., Wang, L., Feng, J., Cao, D., & Pang, B. (2022). Classifying cervical histopathological whole slide images via deep multi-instance transfer learning. In 2022 ieee international conference on bioinformatics and biomedicine (bibm) (pp. 2302–2307).

Karasikov, M., van Doorn, J., Känzig, N., Cesur, M. E., Horlings, H. M., Berke, R., . . . Otálora, S. (2025, October). Training state-of-the-art pathology foundation models with orders of magnitude less data. In Medical image computing and computer assisted intervention – miccai 2025 (Vol. LNCS 15967, pp. 573–583). Springer Nature Switzerland. doi: 10.1007/978-3-032-04984-1\_55

Kingma, D. P., & Ba, J. (2014). Adam: A method for stochastic optimization. arXiv preprint arXiv:1412.6980 .

Li, T., Feng, M., Wang, Y., & Xu, K. (2021). Whole slide images based cervical cancer classification using self-supervised learning and multiple instance learning. In 2021 ieee 2nd international conference on big data, artificial intelligence and internet of things engineering (icbaie) (pp. 192–195).

Mohammadi, M., Fell, C., Morrison, D., Bell, S., Bryson, G., Syed, S., . . . others (2025). Cervical whole-slide images dataset for multiclass classification. GigaScience, 14, giaf144.

Mohammadi, M., Fell, C., Morrison, D., Syed, S., Konanahalli, P., Bell, S., . . . Harris-Birtill, D. (2024). Automated reporting of cervical biopsies using artificial intelligence. PLOS Digital Health, 3 (4), e0000381.

Mudenda, V., Malyangu, E., Sayed, S., & Fleming, K. (2020). Addressing the shortage of pathologists in africa: Creation of a mmed programme in pathology in zambia. African Journal of Laboratory Medicine, 9 (1), 1–7.

Nechaev, D., Pchelnikov, A., & Ivanova, E. (2024). Hibou: A family of foundational vision transformers for pathology. Retrieved from https://arxiv.org/abs/2406.05074

Neidlinger, P., El Nahhas, O. S., Muti, H. S., Lenz, T., Hoffmeister, M., Brenner, H., . . . others (2025). Benchmarking foundation models as feature extractors for weakly supervised computational pathology. Nature biomedical engineering, 1–11.

Nelson, A. M., Milner, D. A., Rebbeck, T. R., & Iliyasu, Y. (2016). Oncologic care and pathology resources in africa: survey and recommendations. Journal of Clinical Oncology, 34 (1), 20–26.

Oliveira, S. P., Montezuma, D., Moreira, A., Oliveira, D., Neto, P. C., Monteiro, A., . . . others (2023). A cad system for automatic dysplasia grading on h&e cervical whole-slide images. Scientific Reports, 13 (1), 3970.

Pal, A., Xue, Z., Desai, K., Banjo, A. A. F., Adepiti, C. A., Long, L. R., . . . Antani, S. (2021). Deep multiple-instance learning for abnormal cell detection in cervical histopathology images. Computers in Biology and Medicine, 138, 104890.

Pina, A., Lavallée, S., Ndiaye, C., & Mayrand, M.-H. (2013). Reproductive impact of cervical conization. Current Obstetrics and Gynecology Reports, 2 (2), 94–101.

Saillard, C., Jenatton, R., Llinares-López, F., Mariet, Z., Cahané, D., Durand, E., & Vert, J.-P. (2024). H-optimus-0. Retrieved from https://github.com/bioptimus/releases/tree/main/models/h-optimus/v0

Skokowski, J., Bolcewicz, M., Jendernalik, K., Vanelslander, T., Gulczyński, J., Lewandowska, A., & Kalinowski, L. (2022). The digital tissue and cell atlas and the virtual microscope.

Sornapudi, S., Addanki, R., Stanley, R. J., Stoecker, W. V., Long, R., Zuna, R., . . . Antani, S. (2021). Automated cervical digitized histology whole-slide image analysis toolbox. Journal of Pathology Informatics, 12 (1), 26.

Tranbaloc, P. (2002). Métaplasie et cin de haut grade. difficultés diagnostiques. Gynécologie obstétrique & fertilité, 30 (11), 845–849.

van der Walt, S., Schönberger, J. L., Nunez-Iglesias, J., Boulogne, F., Warner, J. D., Yager, N., . . . the scikit-image contributors (2014). scikit-image: image processing in python. PeerJ, 2, e453. doi: 10.7717/peerj.453

Vorontsov, E., Bozkurt, A., Casson, A., Shaikovski, G., Zelechowski, M., Liu, S., . . . Fuchs, T. J. (2024). Virchow: A million-slide digital pathology foundation model. Retrieved from https://arxiv.org/abs/2309.07778

Wang, X., Cai, D., Yang, S., Cui, Y., Zhu, J., Wang, K., & Zhao, J. (2023). Sac-net: Enhancing spatiotemporal aggregation in cervical histological image classification via label-efficient weakly supervised learning. IEEE Transactions on Circuits and Systems for Video Technology .

Weinstein, J. N., Collisson, E. A., Mills, G. B., Shaw, K. R., Ozenberger, B. A., Ellrott, K., . . . Stuart, J. M. (2013). The cancer genome atlas pan-cancer analysis project. Nature genetics, 45 (10), 1113–1120.

Zhai, X., Kolesnikov, A., Houlsby, N., & Beyer, L. (2022). Scaling vision transformers. In Proceedings of the ieee/cvf conference on computer vision and pattern recognition (pp. 12104–12113).

Zhao, M., Ling, M., Wang, Z., Shi, J., Kan, H., An, H., . . . Lu, W. (2022). Whole slide image multi-classification of cervical epithelial lesions based on unsupervised pre-training. In 2022 44th annual international conference of the ieee engineering in medicine & biology society (embc) (pp. 594–598).

